# Individual-Level Heterogeneity in Mask-wearing During the COVID-19 Pandemic in Malaysia

**DOI:** 10.1101/2021.07.02.21257837

**Authors:** Stephen X. Zhang, Kim Hoe Looi, Nicolas Li, Xue Wan, Jizhen Li

## Abstract

Wearing a face mask has been a key approach to contain or slow down the spread of COVID-19 in the ongoing pandemic. However, there is huge heterogeneity among individuals in their willingness to wear face masks during an epidemic. This research aims to investigate the individual heterogeneity to wear face masks and its associated predictors during the COVID-19 pandemic when mask-wearing was not mandatory but individuals’ choices. Based on a survey of 708 Malaysian adults and a multivariate least-squares fitting analysis, the results reveal a significant variance among individuals in wearing masks, as 34% of the individual adults did not always wear masks in public places. Female, individuals who wash their hands more frequently, and those who reported more availability of personal protective equipment were more likely to practice mask-wearing. The identification of less compliant groups of mask-wearing has critical implications by enabling more specific health communication campaigns.

---

Since the WHO declared COVID-19 a pandemic, the primary non-pharmaceutical containments at the individual level includes wearing face masks, improved hygiene, and other physical barriers). Health agencies worldwide recommended the compliance of wearing face masks in public during the COVID-19 pandemic. For example, the US Centers for Disease Control and Prevention (CDC) recommended: “ Cover your mouth and nose with a mask when around others”. ^1^ WHO recommended medical/surgical face masks as a key measure of infection prevention and control against the transmission of COVID-19.^2, 3^ Epidemiological modeling results suggest a potentially high value of wearing face masks by the general public to curtail community transmission of COVID-19.^5^ Despite an increasing number of healthcare campaigns to urge community-wide face mask-wearing^3, 4, 6-9^, individual compliance towards face mask-wearing remains problematic,^4,5^ and mask-wearing could present extra challenges for introducing extra stuffy and sweaty feelings in the tropical climate.

Scholars have noticed different receptiveness to face mask-wearing across countries.^9^ For example, in East Asia, face mask-wearing can be quite ubiquitous and was easy to become mandatory quickly in the pandemic.^6-8, 10^ In many countries, many people are reluctant and/or oppose wearing face masks, regarding the wearing of face masks as a symbol of individual freedom.^6-8^ However, to date there remains a void on research that assesses which subgroups of the general population are more or less likely to wear face masks. Research on the individual-level heterogeneity in face mask-wearing within the same country is especially important because such research can help identify the less compliant groups under the same policy to enable the deployment of more targeted health information campaigns on mask-wearing during the ongoing COVID-19 crisis and future pandemics. Under the prolonged COVID-19 pandemic, we need evidence-based research to generate more targeted actions to get the most out of the thin resources compared to the scale of the pandemic, and hence the identification of the risk factors of mask-wearing non-compliance behaviors becomes even more relevant and important.

To date, there have been only four publications on the individual heterogeneity to wear masks, and all the papers are distinct from our study in their foci ^11-14^. Among the four publications, Hao et al. (2021) used state-level variables such as the death rate across 10 states in the USA to predict individuals’ mask-wearing^11^. Kim et al. (2020) and Barcelo & Sheen (2020) studied the antecedents of mask-wearing by the social norms in Korea and Spain respectively.^12,13^ Cherry et al. (2021) ran an online experiment with participants from an M-Turk panel to identify public health messaging and personal experience on the acceptance of mask-wearing ^14^. None of them had the same focus as our study. Furthermore, the four studies were in the USA, Korea, and Spain, and our setting of Malaysia represents the first country in the tropics with a much warmer climate, which may be relevant to mask-wearing as well.

To that end, this study investigates individual adults’ levels of face mask-wearing behavior during the COVID-19 pandemic and explores several predictors based on demographic factors and hygienic factors.

We conducted a survey in May 2020, before Malaysia implemented mandatory mask-wearing in August 2020. The Malaysian Ministry of Health has been advocating wearing face masks as a complementary personal protective device against COVID-19 and has conducted many broad health campaigns with face mask-wearing as an essential element. ^15^ However, face mask-wearing was not mandatory at the time of the study.

Following previous studies^11-14^, we conducted a cross-sectional design because almost all the research on the risk factors is cross-sectional during the COVID-19 crisis,^16^ as the COVID-19 crisis makes it very challenging to pursue other designs. The questionnaire is available in all three major languages (i.e., Malay, Mandarin, and English) in Malaysia. The cross-sectional survey was implemented online, which was a safe and feasible way of collecting data during the pandemic, by a two-stage stratified sampling in terms of geographical area, ethnicity, gender, and age, in line with other similar studies. To minimize biases, we followed the standard survey approaches, that is, no social pressure to influence responses, no questions that provoke defensiveness or threaten esteem, no payoff or cost for particular responses. To avoid common method bias, multi-item questions were used to ensure no priming, and there was no overlapping among questions for different constructs. Participation in this survey was voluntary, and respondents could opt out at any time. Moreover, respondents were assured anonymity and confidentiality of their responses. All respondents consented to the survey, which was ethically approved (#20200322).

The list of variables collected is shown below and the detailed scales are in the online appendix.

- *Gender:* 1 for male and 2 for female.
- *Age:* ranged from 21 years old to 71 years old.
- *The number of children in the household:* from 1 (none) to 7 (six or more children).
- *Handwashing:* Handwashing was measured from 1 (never) to 7 (every time).
- *Availability of PPE*. Given the availability of PPE (personal protective equipment) was an issue early on in the pandemic, the respondents reported the extent to which they had sufficient personal protective equipment PPE from 1 (never) to 5 (always).
- *Face mask-wearing*. The respondents reported how frequently they wore facemasks when outside of their residence from 1 (never) to 7 (every time).

The survey yielded 708 valid responses. We used SPSS (v.26) to run a multivariate least-square regression at the significance level of 0.05.

Table 1 shows the descriptive statistics of the sample (the distribution, mean and standard deviation of the Likert scales). Overall, this study found a higher level of face mask compliance, even though the data for this study were collected before the mandatory face mask-wearing in public places. About two-thirds (66.0%) of the adults wore masks all the time when outing, and the rest 33.4% wore them to varying frequencies. A tiny percentage (0.5%) never wore masks under the pandemic.

**Table 1.** Descriptive statistics of respondents

| Variables | Frequency (%) |  | Mean (S.D.) |  |
| --- | --- | --- | --- | --- |
| **Gender** |  |  |  |  |
| Male | 345 (48.7%) |  |  |  |
| Female | 363 (51.3%) |  |  |  |
| **Age bracket** |  |  |  |  |
| 19 to 29 years old | 101 (14.3%) |  |  |  |
| 30 to 39 years old | 213 (30.1%) |  |  |  |
| 40 to 49 years old | 200 (28.2%) |  |  |  |
| 50 to 59 years old | 159 (22.5%) |  |  |  |
| 60 years old or above | 35 (4.9%) |  |  |  |
| **Number of children in the household** |  |  |  |  |
| 0 | 335 (47.3%) |  |  |  |
| 1 | 119 (16.8%) |  |  |  |
| 2 | 112 | (15.8%) |  |  |
| 3 | 83 | (11.7%) |  |  |
| 4 | 41 | (5.8%) |  |  |
| 5 | 14 | (2.05%) |  |  |
| 6 or more | 4 | (.6%) |  |  |
| **Handwashing** |  |  |  |  |
| Never | 3 | (.4%) |  |  |
| Rarely (less than 10% of the time) | 17 | (2.4%) |  |  |
| Occasionally (about 30% of the time) | 60 | (8.5%) |  |  |
| Sometimes (about 50% of the time) | 106 | (15.0%) | 5.3 | (1.4) |
| Frequently (about 70% of the time) | 199 | (28.1%) |  |  |
| Usually (about 90% of the time) | 119 | (16.8%) |  |  |
| Every time | 204 | (28.8%) |  |  |
| **Availability of PPE** |  |  |  |  |
| Never | 7 | (1.0%) |  |  |
| Rarely | 11 | (1.6%) |  |  |
| Sometimes | 57 | (8.1%) | 4.5 | (.8) |
| Often | 170 | (24.0%) |  |  |
| Always | 463 | (65.4%) |  |  |
| **Face mask-wearing** |  |  |  |  |
| Never | 4 | (.6%) |  |  |
| Rarely (less than 10% of the time) | 8 | (1.1%) |  |  |
| Occasionally (about 30% of the time) | 15 | (2.1%) |  |  |
| Sometimes (about 50% of the time) | 27 | (3.8%) | 6.3 | (1.2) |
| Frequently (about 70% of the time) | 83 | (11.7%) |  |  |
| Usually (about 90% of the time) | 104 | (14.7%) |  |  |
| Every time | 467 | (66.0%) |  |  |

Table 2 shows the regression result to predict individual adults’ face mask-wearing. Among the demographic factors, gender positively predicted face mask-wearing, but not age and the number of children in the household did not. In terms of personal hygiene factors, both handwashing and the availability of PPE positively predicted face mask-wearing. Furthermore, as a robustness test, we have added additional controls such as the education level, ethnicity, income, etc. and the findings remain robust.

**Table 2.** Predictors of face mask-wearing among 708 individual adults

| Variables | Standardized<br>coefficient | t | p-value | 95% confidence interval |  |
| --- | --- | --- | --- | --- | --- |
|  |  |  |  | Lower<br>Bound | Upper<br>Bound |
| Gender (female) | .078 | 2.339 | .020 | .029 | .333 |
| Age bracket | .017 | .499 | .618 | -.052 | .088 |
| Number of children in<br>household | -.042 | -1.261 | .208 | -.088 | .019 |
| Handwashing frequency | .156 | 4.355 | .000 | .070 | .186 |
| PPE availability | .395 | 11.273 | .000 | .477 | .678 |

Our finding on gender is consistent with recent studies on the gender difference in terms of face mask-wearing. A US-based study found that female shoppers were 1.5 times more likely to wear face masks than male shoppers. ^17^ And a study in Saudi Arabia revealed the same gender difference.^18^ Our results reveal that two key individual-level hygienic factors during epidemics – handwashing and PPE availability– predicted face mask-wearing. Our results demonstrate those who were less likely to practice handwashing or had less PPE available were also less unlikely to practice face mask-wearing. The identification of these predictors enables healthcare organizations to identify statistically the less compliant groups to enable the deployment of more targeted health information campaigns on mask-wearing during the ongoing COVID-19 crisis and future pandemics. Such identification of the less compliant groups enables more targeted actions to get the most out of the thin resources compared to the scale of the pandemic, and hence the identification of the risk factors of mask-wearing non-compliance behaviors is crucial in the prolonged pandemic.

This research is not without limitations. To begin with, our survey captured the individual heterogeneity at a single point in the middle of the pandemic. It would be ideal for learning how individuals’ mask-wearing behaviors change as the pandemic continues to develop. In terms of methodology, the web-based design means that people with no internet access and limited computer literacy were not surveyed, which made it hard for us to the older population. Our study used a cross-sectional survey which is popular in identification studies in COVID-19 literature, and future studies may use matched or control case datasets. Moreover, other predictors, such as the need to go out, the locations individuals venture out, as well as psychological factors including but not limited to their individual perception^19^, perceived risk^20^, knowledge, perceived benefit, trust, social media, etc. can be studied as additional predictors of mask-wearing. Last but not least, given the measure of the dependent variable, we used linear regression instead of logistic regression. We call future research to study mask-wearing not only by its frequency with linear regression but also by yes or no with logistic regression.

In conclusion, by reporting the heterogeneity of individual adults’ face mask-wearing and identifying several associated predictors, this study advanced research on mask-wearing. The results revealed a huge individual heterogeneity in wearing masks and found individual’ s gender, hand washing, and the availability of personal protective equipment predicted their face mask-wearing. Practically, this evidence-based research can help public health organizations to identify the less compliant groups to enable the deployment of more targeted health information campaigns on mask-wearing during the ongoing COVID-19 crisis and future pandemics.

## Data Availability

available upon request

## Author Contributions

- conceptualization, S. X. Z.; K. H. L.; N. L. J. L.;
- methodology, S. X. Z.; K. H. L.; N. L.
- software, S. X. Z.; K. H. L.; N. L.
- validation, S. X. Z.; K. H. L.; N. L. X. W.
- formal analysis, S. X. Z.; K. H. L.; N. L.
- the investigation, S. X. Z.; K. H. L.; N. L. X. W.
- resources, J. L.
- data curation, K. H. L.
- writing—original draft preparation, S. X. Z.; K. H. L.; N. L.
- writing—review and editing, S. X. Z.; K. H. L.; N. L. X. W.
- visualization, K. H. L.; N. L.
- supervision, S. X. Z.; K. H.
- project administration, S. X. Z.; K. H.
- funding acquisition, J. L.;

All authors have read and agreed to the published version of the manuscript.

## Funding

We acknowledge the support from the National Natural Science Foundation of China [grant number 71772103].

## Institutional Review Board Statement

The study was conducted according to the guidelines of the Declaration of Helsinki, and approved by the Institutional Review Board (or Ethics Committee) of Tsinghua University (20200322).

## Informed Consent Statement

Informed consent was obtained from all subjects involved in the study.

## Data Availability Statement

Data are available upon request

## Conflicts of Interest

The authors declare no conflict of interest.

## Appendix: the scale of face mask-wearing behavior

How frequent do you wear a face mask when you go outside of your house?/

a. Never
b. Rarely (less than 10% of the time)
c. Occasionally (about 30% of the time)
d. Sometimes (about 50% of the time)
e. Frequently (about 70% of the time
f. Usually (about 90% of the time)
g. Every time

## Notes

### Competing Interest Statement

The authors have declared no competing interest.

### Author Declarations

Tsinghua University (ethical approval #: 20200322)

